# Optimizing SARS-CoV-2 vaccination strategies in France: Results from a stochastic agent-based model

**DOI:** 10.1101/2021.01.17.21249970

**Authors:** Nicolas Hoertel, Martin Blachier, Frédéric Limosin, Marina Sánchez-Rico, Carlos Blanco, Mark Olfson, Stéphane Luchini, Michaël Schwarzinger, Henri Leleu

## Abstract

The COVID-19 pandemic is a major global societal, economic and health threat. The availability of COVID-19 vaccines has raised hopes for a decline in the pandemic. We built upon a stochastic agent-based microsimulation model of the COVID-19 epidemic in France. We examined the potential impact of different vaccination strategies, defined according to the age, medical conditions, and expected vaccination acceptance of the target non-immunized adult population, on disease cumulative incidence, mortality, and number of hospital admissions. Specifically, we examined whether these vaccination strategies would allow to lift all non-pharmacological interventions (NPIs), based on a sufficiently low cumulative mortality and number of hospital admissions. While vaccinating the full adult non-immunized population, if performed immediately, would be highly effective in reducing incidence, mortality and hospital-bed occupancy, and would allow discontinuing all NPIs, this strategy would require a large number of vaccine doses. Vaccinating only adults at higher risk for severe SARS-CoV-2 infection, i.e. those aged over 65 years or with medical conditions, would be insufficient to lift NPIs. Immediately vaccinating only adults aged over 45 years, or only adults aged over 55 years with mandatory vaccination of those aged over 65 years, would enable lifting all NPIs with a substantially lower number of vaccine doses, particularly with the latter vaccination strategy. Benefits of these strategies would be markedly reduced if the vaccination was delayed, was less effective than expected on virus transmission or in preventing COVID-19 among older adults, or was not widely accepted.

## Introduction

The COVID-19 pandemic is a major global societal, economic and health threat. In the initial absence of a vaccine or an effective treatment for COVID-19, most countries have responded with a variety of non-pharmaceutical interventions (NPIs), such as physical distancing, mandatory use of face masks, and repeated lockdowns, to curtail viral transmission by reducing contact rates and to avoid the overwhelming of healthcare services. These measures have been effective in reducing disease transmission, but have led to negative psychological ^1–3^, medical ^4,5^, economic, and social consequences. ^6,7^

Recent phase III trials have demonstrated that three vaccines could prevent development of severe form of SARS-CoV-2 infection and reduce disease transmission, raising hopes for a decline of the pandemic.^8–10^ However, beyond vaccine efficacy, the success of a vaccine depends on the vaccination program strategy, including the identification of the priority target population, its availability timelines constrained by the production and distribution capabilities, and its rates of uptake in the target population. In the context of limited supply of COVID-19 vaccines, there is a need to define priority groups within the populations. In France, COVID-19 vaccine acceptance in the general population ranges from 50% to 60%, with substantial variability across age groups.^11–14^ Therefore, determining vaccination program strategies that optimally mitigate the pandemic according to these parameters is urgently needed.

In this report, we built upon a stochastic agent-based microsimulation (ABM) model of the COVID-19 epidemic in France,^15^ and projected the potential impact of different vaccination strategies on COVID-19 cumulative incidence, mortality, and number of hospital admissions. Specifically, because NPIs impose considerable burdens on the population and the economy, we examined whether prioritizing vaccination of older adults or individuals with medical conditions, who are the most prone to develop severe COVID-19,^16^ would permit lifting NPIs, based on a sufficiently low cumulative mortality and number of hospital admissions.

## Methods

### Model structure

We built upon a stochastic agent-based model of the epidemic of COVID-19 in France that previously showed adequate calibration and validation.^15^ Briefly, this model includes (i) a realistic synthetic population generated with demographic characteristics, medical comorbidities and household structure representative of the French general population,^17–20^ (ii) a social contact network among the individuals, each with a geolocalized activity sequence over the day, taking into account co-location probability and duration, including contacts with family members, extended family members or friends (at home or at bars and restaurants), contacts at school or at work, and during public transport or grocery shopping or cultural activities, and (iii) a disease model, which translates the edge weights in the social contact network into infection probability of the edge over the day. The model parameters are summarized in **eTable 1**. We updated the contamination risk and proportion of undiagnosed cases of our initial model^15,21^ and included data on SARS-CoV-2 seroprevalence in May 2020 in France.^22^

### Outcomes

Outcomes included cumulative incidence, mortality, and number of hospital admissions.

The probabilities of hospital admission or death were stratified by age and adjusted for comorbidities, including, obesity, diabetes, chronic cardiac diseases, and chronic respiratory diseases, based on hazard ratios calculated using data from Institut Pasteur^23^ and from the OpenSAFELY cohort study.^16^ To reflect improved care of patients with COVID-19, we reduced the risk of death by an average of 10% in the model, regardless of age, starting July 1, 2020 to fit observational data.^24,25^ Delays between infection, symptom onset, hospital admission, death or recovery were based on prior reports.^23,26,27^

### Vaccine Efficacy

COVID-19 vaccine efficacy was assessed using published results for the BNT162b2 mRNA COVID-19 Vaccine from Pfizer/BioNTech.^10^ Based on these data, the efficacy of two doses of the BNT162b2 mRNA COVID-19 Vaccine is expected to be 95.6% (95% CI: 89.4%–98.6%) in individuals aged 16 to 55 years, 93.7% (95% CI: 80.6%–98.8%) in those aged 55 to 65 years, and 94.7% (95% CI: 66.7%–99.9%) in those aged 65 to 75 years. For individuals aged over 75 years, we assumed a similar efficacy as in those aged 65 to 75 years. The efficacy of two doses of COVID-19 Vaccine mRNA-1273 from Moderna^9^ is expected to be very similar, with an estimated rate of 94.5% (95% CI: 86.5%-97.8%) in individuals aged 18 years and over. Finally, because the efficacy reported for two doses of the ChAdOx1 nCoV-19 vaccine from AstraZeneca/Oxford^8^ was substantially lower, i.e., 70.4% (95% CI: 54.8%-80.6%), in individuals aged 18 years and over, we conducted sensitivity analyses using efficacy data of this vaccine.

### Vaccine uptake

Estimated COVID-19 vaccine acceptance was based on a discrete choice experiment conducted in a large sample representative of the French population aged 18-64 years.^11^ This study showed that vaccine uptake would be expected to assume an inverted U-shape relationship with advancing age. We assumed in our model that individuals accepting the vaccine would be vaccinated. In the absence of specific data, we also assumed that vaccine acceptance in the population aged 65 years and over would be similar to that reported for individuals aged 55 to 64 years.

### Statistical Analysis

The stochastic agent-based microsimulation model of the COVID-19 epidemic in France was run using C++ from March 1^st^, 2020, until August 1^st^, 2021, on 500,000 individuals with an average of 200 simulations. The results were extrapolated to the French population, which comprises about 67 million people. We provided uncertainty measures by using 100 bootstrap samples based on the random variation of all parameters simultaneously, excluding vaccination acceptance to facilitate interpretation, either within their 95% confidence interval for parameters estimated from the literature or within a +/- 20% interval if the parameter was assumed.^15,21^ All results are presented per 100,000 inhabitants to facilitate international comparisons.

We examined whether the model had adequate calibration, i.e., whether it was able to adequately reproduce retrospectively the course of the epidemic until December 20^th^, 2020, based on R^2^ and Normalized Root Mean Squared Error (NRMSE) for weekly mortality and hospital admissions, and visual comparison between model-predicted and observed mortality and hospital admissions.

All scenarios included a full population lockdown between March 17^th^, 2020 and May 11^th,^ 2020, followed by a progressive return to 75% of the pre-pandemic social contacts level until July 1^st^, 2020, except at schools, which remained closed during that period, and a 30% rate of workers using telework. Following prior epidemiological trends,^28^ we assumed that SARS-CoV-2 infection was associated with a 23% reduction in disease transmission due to warm weather between July 1^st^, 2020 and September 25^th^, 2020. This latter date was chosen because it marked a significant drop in temperature in France and was quickly followed by a significant increase in the number of cases. On September 1^st^, 2020, schools reopened for all students and telework use decreased to 16% based on Google Mobility Reports for France.^24^ Based on data from Santé Publique France,^24^ we considered that face mask use at work, in public transport, during grocery shopping and for cultural events increased from 15% to 70% between April 4^th^ and September 1^st^, 2020, and remained at this level hereafter. We assumed a limited use of face mask (i.e., 30%) in households or with friends or extended family members during this period and hereafter. Curfew was instated on October 17^th^, 2020, which has led to cancelation of all cultural events and was assumed to reduce social contacts with friends and extended family members by 50%. We considered that this curfew would last until January 15^th^, 2021. A second less stringent lockdown was instated between October 30^th^ and December 15^th^, 2020, with schools and workplaces remaining opened. We assumed that 50% of individuals worked remotely from home during this second lockdown period and that this rate will be of 30% during the curfew after the second lockdown lifting.

To examine whether any vaccination scenario could allow for a lifting of NPIs, we assumed that from January 15^th^, 2021, social behaviors would return to those observed before the COVID-19 epidemic, with full discontinuation of all NPIs, and examined associated cumulative incidence, mortality, and number of hospital admissions. In our model, we considered that a vaccination strategy would allow for the discontinuation of NPIs if it was associated with (i) a cumulative number of deaths lower than 17 per 100,000 and (ii) a cumulative number of hospital admissions below 240 per 100,000, between December 27^th^, 2020 and August 1^st^, 2021. The first threshold corresponds to the mean plus two standard deviations of the total number of deaths observed in France between January 15^th^, 2020 and August 1^st^ for the years 2015 to 2019, representing the threshold above which an increase in death could be considered significant. Given that hospital-bed capacity is 600 per 100,000 inhabitants in France^24^ and that the mean duration of a hospitalization for COVID-19 is about 21 days,^24^ the second threshold corresponds to a maximum hospital-bed occupancy rate for COVID-19 of 5% between December 27^th^, 2020 and August 1^st^, 2021. We chose this threshold because current hospital-bed occupancy by patients with COVID-19 is currently estimated at 6.3% (25,000/400,000) of the total number of hospital beds in France.^24^

Given the limited production and distribution capabilities for COVID-19 vaccines, it is expected that vaccinating the full non-immunized French population aged 18 years or older would probably require several months, even with three vaccines. Because our main aim was to examine whether different vaccination strategies would allow for lifting NPIs, rather than make questionable assumptions on time for vaccinating the population, we considered that vaccination in each scenario would be achieved by January 15^th^, 2021 and calculated the number of vaccine doses needed (considering 2 doses per individual) in each scenario.

Next we examined the impact of different vaccination scenarios according to the choice of the non-immunized adult populations to prioritize for vaccination: (i) no vaccination, (ii) vaccination of the full population, (iii) vaccination of adults aged less than 65 years, (iv) vaccination of adults aged more than 45 years, (v) vaccination of adults aged less than 35 years or more than 65 years, (vi) vaccination of adults aged more than 65 years, (vii) vaccination of adults aged more than 55 years with mandatory vaccination of adults aged more than 65 years (assuming that it would lead to a 90% vaccination rate in this population), and (viii) vaccination of individuals at higher risk for severe SARS-CoV-2 infection (i.e. adults aged more than 65 years and those with medical conditions associated with increased risk of severe COVID-19^16^). Two factors drove the choice of these scenarios: (i) the substantial risk of severe disease in adults aged more than 65 years and in those with medical conditions, and (ii) the higher expected vaccination uptake in younger than in older adults (**Table 1**).

**Table 1.** Estimated age-stratified vaccine acceptance and number of individuals immunized (i.e. approximated as having had COVID-19) on December 27^th^, 2020.

|  | Age Groups |  |  |  |  |  |
| --- | --- | --- | --- | --- | --- | --- |
|  | 18-24 | 25-34 | 35-44 | 45-54 | 55-64 | 65+ |
| Population size, millions | 5.42 | 7.77 | 8.30 | 8.94 | 8.46 | 13.75 |
| Population diagnosed with COVID-19, millions | 0.47 | 0.38 | 0.33 | 0.32 | 0.26 | 0.35 |
| Expected vaccine uptake rate | 68% | 51% | 59% | 62% | 68% | 68% |
| Expected number of individuals to vaccinate, millions | 3.37 | 3.77 | 4.70 | 5.34 | 5.58 | 9.11 |

We performed several sensitivity analyses for the four following scenarios: (i) vaccination of the full adult non-immunized population, (ii) vaccination of adults aged more than 45 years, (iii) vaccination of adults aged more than 55 years with mandatory vaccination of adults aged more than 65 years, and (iv) vaccination of at-risk individuals. First, we considered a 10% lower rate of vaccine uptake than that expected.^11–14^ Second, we examined the impact on our results of a lower efficacy of the vaccine in preventing COVID-19 among individuals aged more than 75 years (i.e. 50% instead of 94.7%),^29^ since very few COVID-19 cases were reported in Polack et al. in this population.^10^ Third, we reproduced the analyses while considering efficacy data of the ChAdOx1 nCoV-19 vaccine instead of the BNT162b2 mRNA COVID-19 vaccine. Fourth, we examined the impact of delaying the vaccination on the course of the epidemic by considering that the target population of each scenario was vaccinated by April 14^th^ instead of January 15^th^. In this scenario, we assumed that NPIs present on December 15^th^ would be maintained until April 15^th^ and then discontinued after that date. Fifth, given the uncertainty of the effect of the vaccine on virus transmission (e.g. data from the ChAdOx1 nCoV-19 vaccine^8^ suggest an efficacy on carriage that is 60% lower than the immune response, as previously seen in other vaccines,^28,30–32^ and data from BNT162b2 mRNA and mRNA-1273 did not include carriage as an endpoint^9,10^), we tested a scenario where the vaccine would only decrease by 50% the virus transmission of vaccinated individuals with immune response, instead of 100% of those with immune response as in the main analyses. Finally, we examined the robustness of our results by evaluating the impact on outcomes of varying simultaneously all individual parameter values by ±20% for the scenario ‘vaccination of the full adult non-immunized population’.

## Results

### Model calibration

The model calibrated well, based on a good visual fit between observed and model-predicted hospital admissions and mortality (**Figure 1**). In addition, R^2^ and NRMSE were 0.96 and 5.6% for weekly hospital admissions, and 0.96 and 4.6% for weekly mortality, respectively. Based on our model, we projected that the cumulative COVID-19 incidence in France would be 15.7% [95% CI: 14.0%-18.1%] on December 27^th^, 2020.

**Figure 1.**
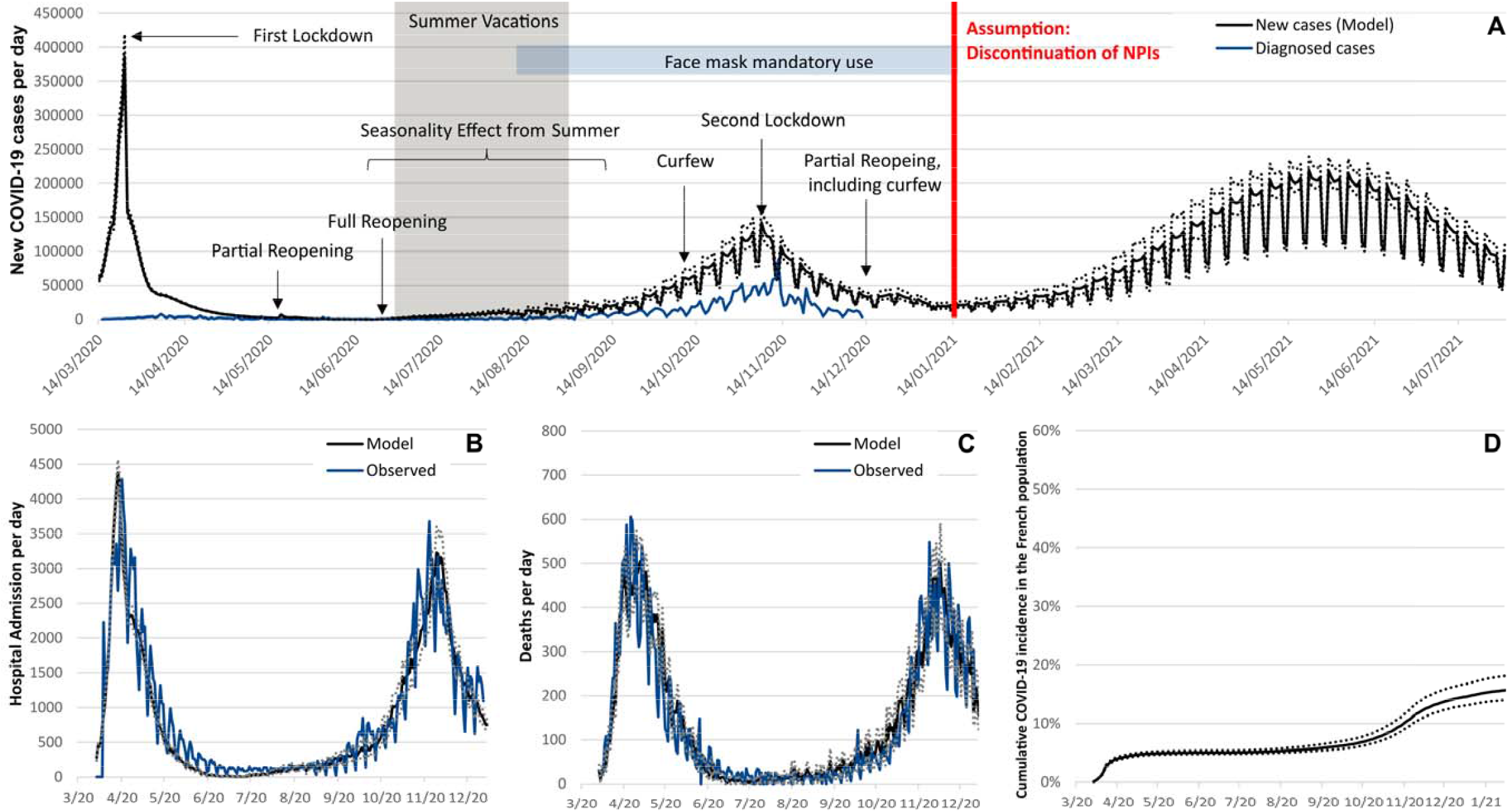
Model-predicted and observed daily incidence (A), number of hospital admissions (B), and mortality (C) related to COVID-19 in France, and model-predicted cumulative incidence of SARS-COV-2 infection (D). The dotted lines represent the uncertainty range (95% prediction interval) stemming from the uncertainty in the parameter values.

### Main analyses

In the absence of a vaccine and in case of lifting all NPIs on December 27^th^, 2020, we projected that between that date and August 1^st^, 2021, a substantial rebound of the COVID-19 epidemic would occur, leading to 32,157 new cases per 100,000 inhabitants [95% prediction interval: 30,447-33,867], a rate of new cases per 100,000 people that would exceed that observed between March 1^st^ and December 27^th^ in France (15,630, [95% interval: 14,000-18,077]) (**Figure 1**). Based on our model, the estimated cumulative mortality and number of hospital admissions between December 27^th^ and August 1^st^, 2021 would be 159 deaths per 100,000 people [95% interval: 145-173] and 876 admissions per 100,000 inhabitants [95% interval: 819-933] (**Table 2**).

**Table 2.**
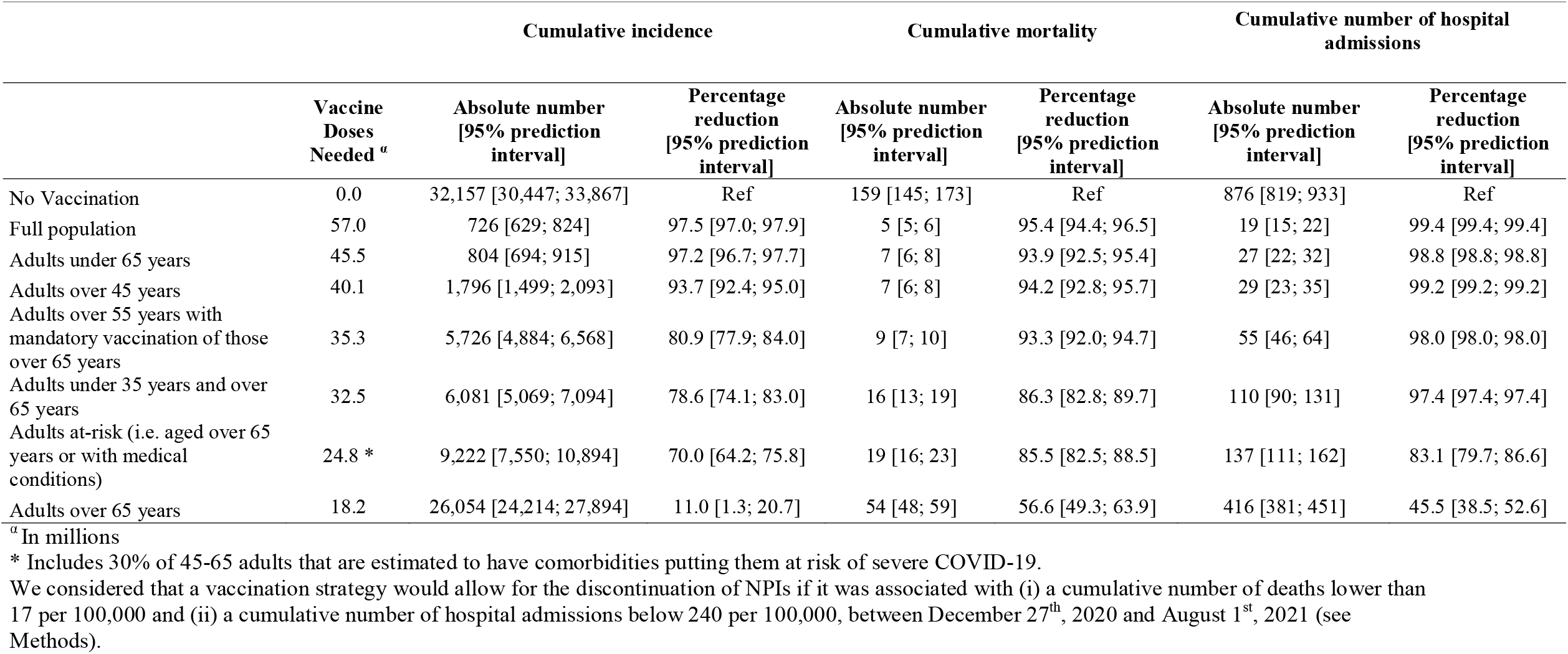
Estimated cumulative incidence, mortality and number of hospital admissions associated with the competing vaccination strategies for non-immunized adults between December 27^th^, 2020 and July 1^st^, 2021.

Immediate vaccination of the full adult non-immunized population, requiring the availability of 57 million vaccine doses (i.e., 2 doses for 28.5 million people), would substantially improve the course of the epidemic as compared to the absence of vaccination, as shown by a 97.5% [95% interval: 97.0; 97.9] decrease in cumulative incidence, a 95.4% [95% interval: 94.4; 96.5] decrease in mortality, and a 97.4% [95% interval: 96.8; 98.0] decrease in cumulative hospital-bed occupancy (**Table 2**).

For scenarios in which the vaccination targeted a specific adult non-immunized population, our findings suggest that all of them, except the scenarios targeting only adults aged more than 65 years, or only at-risk individuals, or only individuals aged over 65 years or under 35 years, would virtually allow lifting of NPIs, if they were applied immediately, based on a cumulative mortality rate lower than 17 per 100,000 and a cumulative hospital admission rate lower than 240 per 100,000 (**Table 2, eFigure 1**). Among the strategies that would allow discontinuing NPIs, i.e., vaccinating only adults under 65 years, or only adults over 45 years, or only adults over 55 years with mandatory vaccination of those over 65 years, the number of vaccine doses needed would be 45.5 million, 40.1 million, and 35.3 million, respectively (**Table 1**).

### Sensitivity analyses (**Table 3**)

A reduction of 10% of the expected rate of vaccine uptake would be associated with a substantial increase in mortality and hospital-bed occupancy, and would prevent discontinuation of NPIs for all strategies, unless the full adult non-immunized population was vaccinated. Lower effect of the vaccine on preventing COVID-19 among individuals aged 75 years or older (i.e. 50% instead of 94.7%) would be associated with a substantially higher cumulative mortality and would prevent the lifting of NPIs in all scenarios, except with the vaccination of the full non-immunized adult population or of the subpopulation aged over 45 years. Similarly, lower vaccine efficacy on carriage (i.e. 50% instead of 100%) would also lead to a cumulative mortality higher than the threshold of 17 per 100,000 for all vaccination strategies, except for those targeting the full non-immunized adult population or the subpopulation of adults aged over 45 years. Delayed vaccination of the target population or a less effective vaccine, as reported for ChAdOx1 nCoV-19, would result in worse outcomes, including higher incidence, mortality, and hospital-bed occupancy, and would not allow for discontinuation of NPIs with current expected rates of vaccination acceptance. Varying all model parameter values by ±20% for the scenario ‘vaccination of the full adult non-immunized population’ would change the cumulative number of hospital admissions by ± 16.2% and the cumulative mortality by ± 18.5%, suggesting the robustness of the differences observed across scenarios.

**Table 3.**
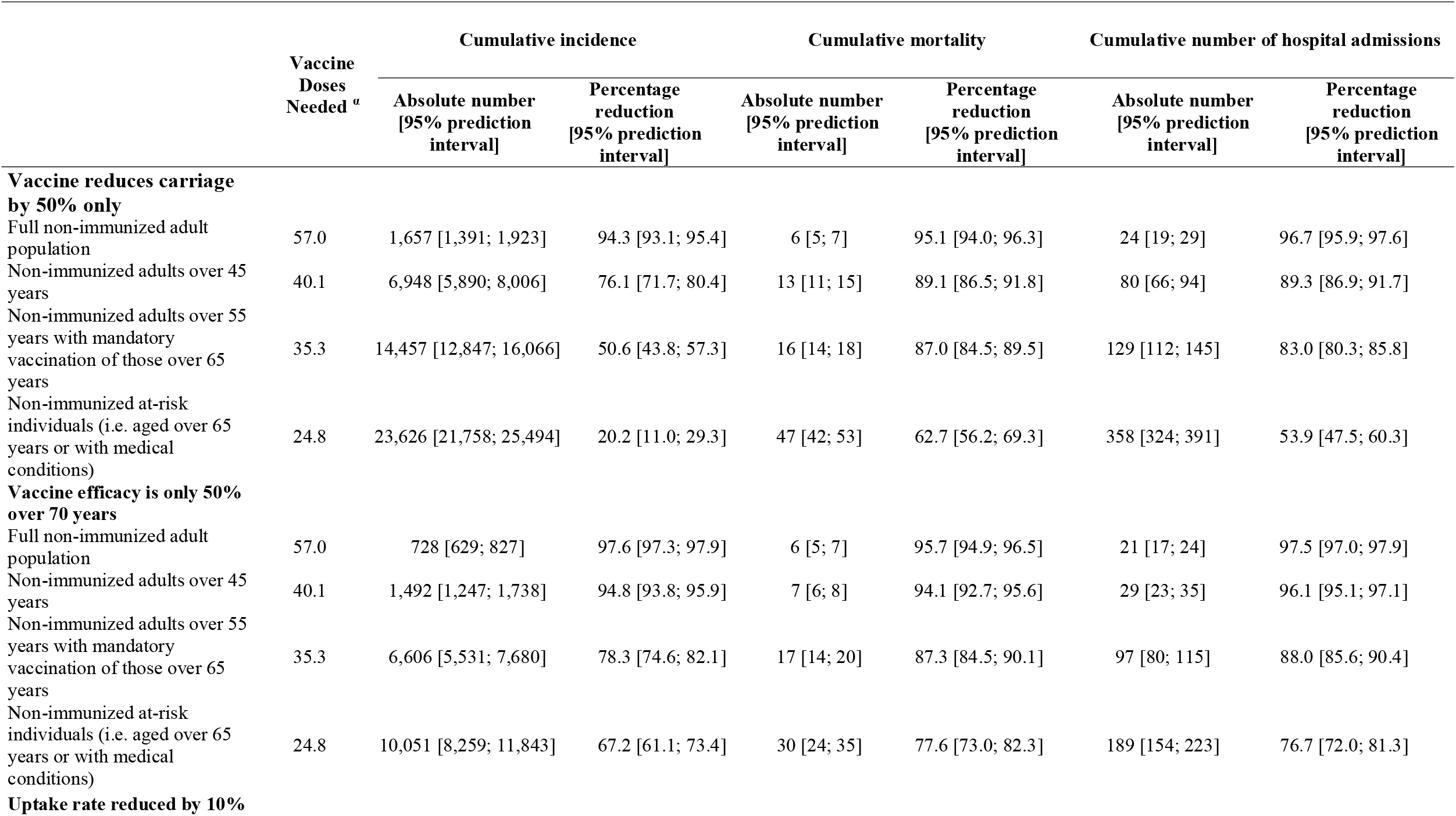
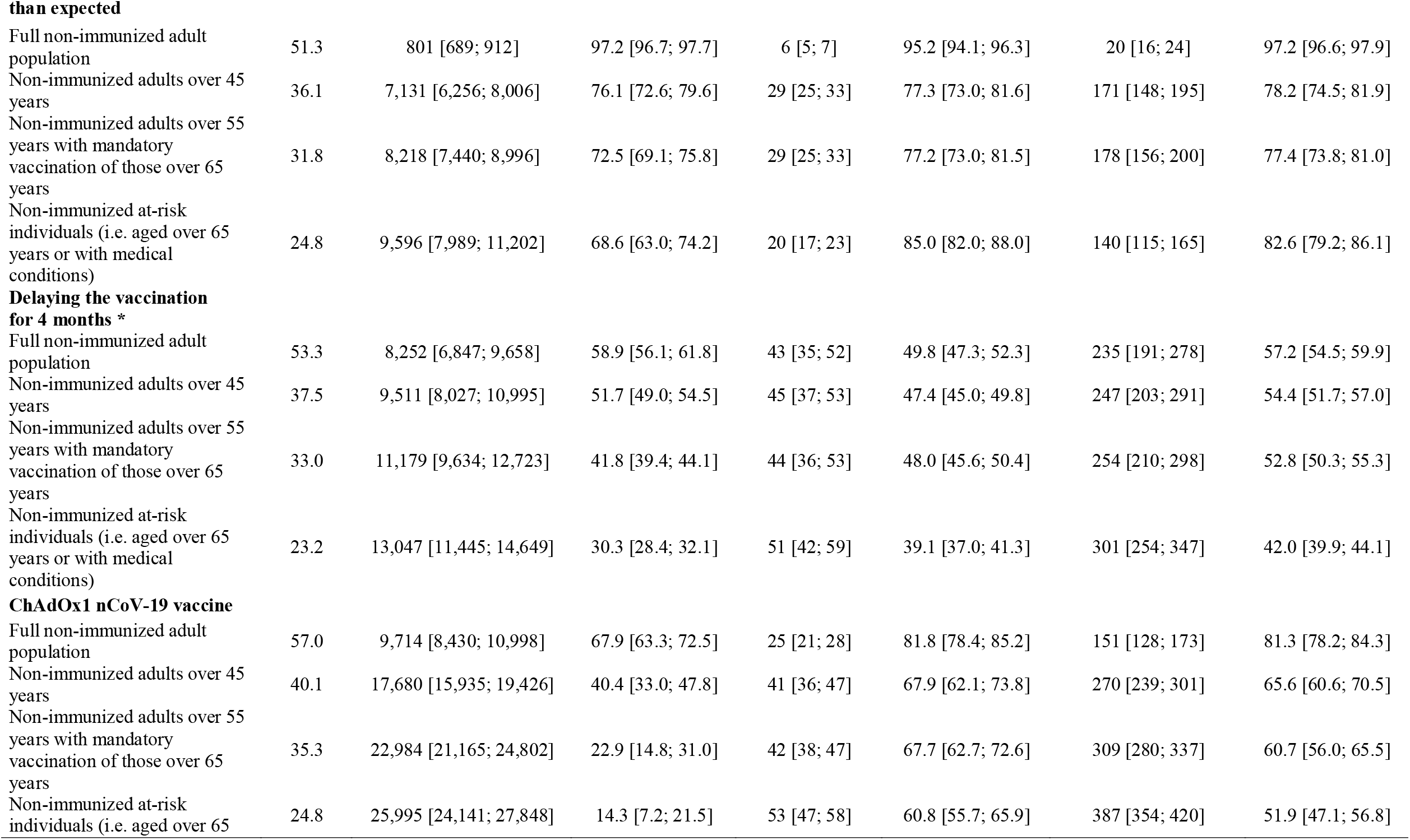
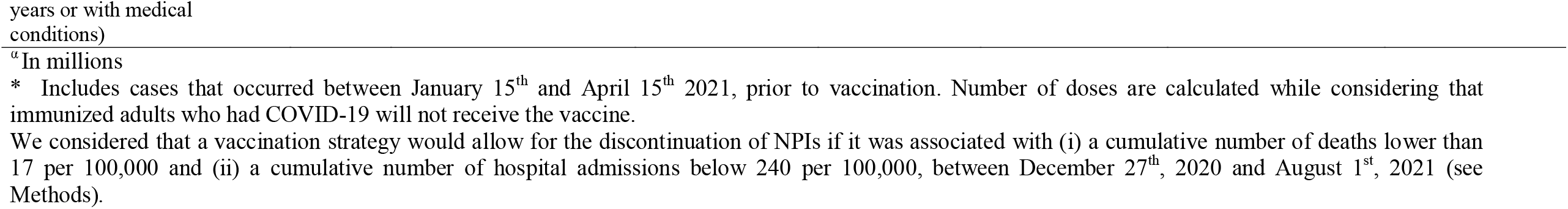
Sensitivity analyses.

## Discussion

In this report, we built upon a stochastic agent-based microsimulation model of the COVID-19 epidemic in France^15^ and examined the potential impact of different vaccination strategies, based on the age, medical conditions, and expected vaccination acceptance of the target non-immunized adult population, on disease cumulative incidence, mortality, and number of hospital admissions. Specifically, we examined whether these vaccination strategies would allow lifting all non-pharmacological interventions, based on a sufficiently low cumulative mortality and number of hospital admissions. The model calibrated well and the variation of each model parameter value by ±20% had limited impact on outcome estimates. While vaccinating the full population, if performed immediately, would be highly effective in reducing incidence, mortality and hospital-bed occupancy, and would allow lifting of all NPIs, this strategy would require 57 million doses of vaccine. Vaccinating only adults aged over 45 years, or only adults aged more than 55 years with mandatory vaccination of those aged over 65 years, would also enable, if performed immediately, lifting all NPIs, but with a substantially lower number of vaccine doses, especially with the latter vaccination strategy (35.3 million doses). Benefits of these strategies would be markedly reduced if the vaccination was delayed, or less effective than expected on virus transmission or in preventing COVID-19, or if most people did not accept vaccination.

Current vaccination plans in France^33^ call for a multi-step approach by decreasing age groups, with priority given for the available vaccine doses to individuals aged over 80 years and healthcare workers. Step 2 of this plan involves vaccinating people over 75 years, then those older than 65 years and having medical conditions increasing the risk of severe SARS-CoV-2 infection,^24^ and finally all other adults between 65 and 74 years of age. While this strategy is likely to markedly reduce mortality and hospital-bed occupancy, ^24^ our results suggest that it would not be sufficient to allow discontinuing NPIs. Even once step 2 is achieved, NPIs would still have to be enforced to prevent a rebound of the epidemic. In our model, an additional 35,235 deaths [95% prediction interval: 31,639; 38,832] in France were projected if NPIs were discontinued after vaccinating only adults aged over 65 years. This result can be explained by two factors. First, the vaccine acceptance in this age group is estimated to be 68%, leaving a substantial fraction of this population at risk of being infected. Second, model results suggest that despite two successive epidemic rebounds, vaccinating only 68% of the individuals aged over 65 years would not be sufficient to produce herd immunity, especially with a high infection rate among younger adults. Indeed, while vaccinating individuals aged over 65 years substantially reduces mortality and the number of hospital admissions compared to the absence of vaccination, it would only reduce by 19% the cumulative COVID-19 incidence. This finding reflects the individual benefit of the vaccination in the absence of herd immunity, with 32% of the population remaining unprotected. Conversely, in a scenario where vaccination coverage rate was 90% for this age group (for example in case of mandatory vaccination), most individuals would benefit from the individual protection provided by a vaccine. Although the scenario of a mandatory vaccination for adults aged over 65 years and a priority given to adults between 55 and 65 years of age may be discussed from an ethical perspective,^33^ our results suggest that it would be associated with a cumulative mortality rate of 9 [95% interval: 7; 10] per 100,000 and a cumulative number of hospital admissions of 55 [95% interval: 46; 64], allowing for discontinuation of NPIs, which are associated with substantial negative psychological,^1–3^ medical,^4,5^ economic, and social consequences^6,7^ in the full population.

There are still some uncertainties regarding the efficacy and the acceptance of COVID-19 vaccines. First, it is not yet known whether vaccine efficacy is similar in individuals aged over 75 years and in younger adults. In our model, lower efficacy among individuals aged 75 years or older (i.e. 50% instead of 94.7%) would be associated with a substantially higher cumulative mortality and would prevent the lifting of NPIs with all vaccination strategies, except those including vaccination of the full non-immunized adult population or of the subpopulation of adults aged over 45 years. Second, estimated acceptance might be lower than expected. Reducing expected vaccine acceptance by 10% did not yield substantially different results when vaccinating the full population, but did make discontinuing NPI less likely with all other scenarios. However, vaccine acceptance might be higher than expected as people learn more about vaccine effectiveness.^34^ Finally, there is debate as to whether vaccines would protect against asymptomatic SARS-CoV-2 carriage,^9^ as clinical trial endpoints were based on COVID-19 diagnosis and not on routine RT-PCR.^8–10^ In our model, lower vaccine efficacy on carriage (i.e. 50% instead of 100%) would lead to a substantially higher cumulative mortality, preventing discontinuation of NPIs in all vaccination strategies, except for those including vaccination of the full non-immunized adult population or of the subpopulation of adults aged over 45 years.

Our study has several limitations. First, as with all modeling studies, we rely on existing knowledge and current assumptions, that might need to be revised with advances in knowledge of this novel disease. Second, there are still uncertainties concerning vaccine effectiveness, availability, and acceptance. Although we used real world data for acceptance and data from large phase III clinical trials for vaccine efficacy, we cannot rule out heterogeneity in vaccine effectiveness and uptake that would be tied to COVID-19 risk. For example, if more at-risk individuals do not accept vaccination, it may reduce the efficacy of the tested strategies.^35^ Third, we considered that infected people could develop immunity for at least several months.^36^ Although post-COVID-19 immunity length remains incompletely known, this assumption has not been rejected, with only a small number of reinfection cases reported. Fourth, we considered that vaccination in each scenario would be virtually achieved by January 15^th^, 2021 and calculated the number of vaccine doses needed in each scenario. While this is unrealistic, our objective was to assess which vaccination strategies might permit safe discontinuation of NPIs if performed immediately. Although implementation of such strategies may require weeks if not months during which NPIs should be maintained, we preferred this approach instead of making uncertain assumptions concerning population behaviors during the next months. Finally, the results should not be interpreted as absolute numbers but rather as differences in expected outcomes according to vaccination strategies.

COVID-19 represents a major public health threat worldwide. The availability of COVID-19 vaccines has raised hopes for a decline of the pandemic. While vaccinating the full adult non-immunized population, if performed immediately, would be highly effective in reducing incidence, mortality and hospital-bed occupancy, and would allow discontinuing all NPIs, this strategy would require a large number of vaccine doses. Vaccinating only adults aged over 45 years or adults aged more than 55 years with mandatory vaccination of those aged over 65 years, would also enable, if performed immediately, lifting of all NPIs with a substantially lower number of vaccine doses, particularly with the latter vaccination strategy. Benefits from these strategies would nonetheless be markedly reduced if the vaccination was delayed, or less effective than expected in preventing virus transmission and COVID-19, or if most people do not accept vaccination.

## Data Availability

Source code of the model has been deposited in a recognized public source code repository (GitHub, https://github.com/henrileleu/covid19).

## Acknowledgments

We thank Pr Melanie Wall and Pr Yuanjia Wang for their helpful comments on an early version of the methods.

## SUPPLEMENTARY MATERIAL

**eFigure 1.**
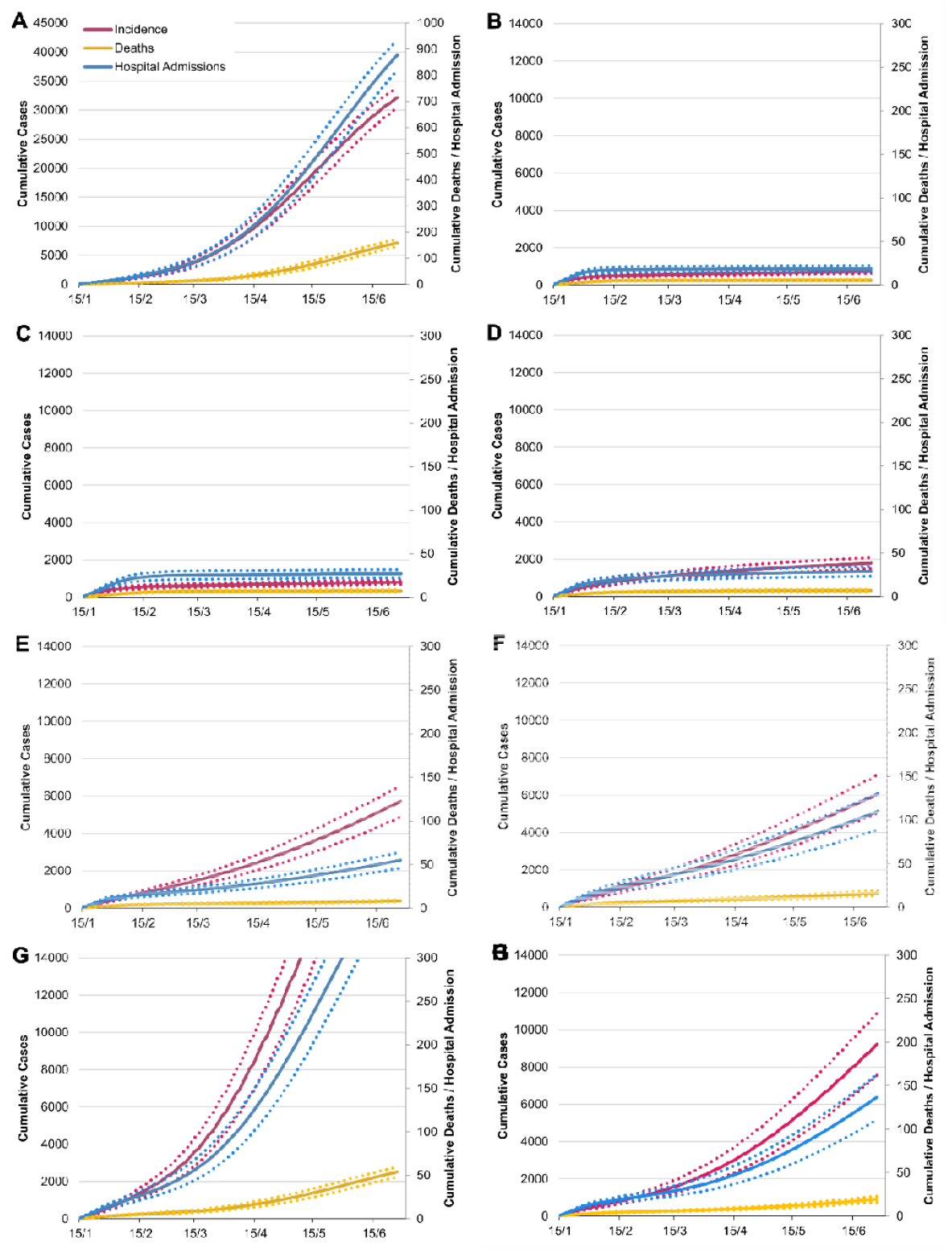
Estimated cumulative incidence, mortality and number of hospital admissions between January 15^th^, 2021 and July 1^st^, 2021 associated with the absence of vaccination (A), and with the vaccination of the non-immunized full adult population (B), adults under 65 years (C), adults over 45 years (D), adults over 55 years with mandatory vaccination of those over 65 years (E), adults under 35 years and over 65 years (F), adults over 65 years (G), and adults at risk of severe COVID (i.e. aged over 65 years or with medical conditions) (H). Note: Numbers are absolute values for the French population. The dotted lines represent the uncertainty range (95% prediction interval) stemming from the uncertainty in the parameter values.

**eFigure 2.**
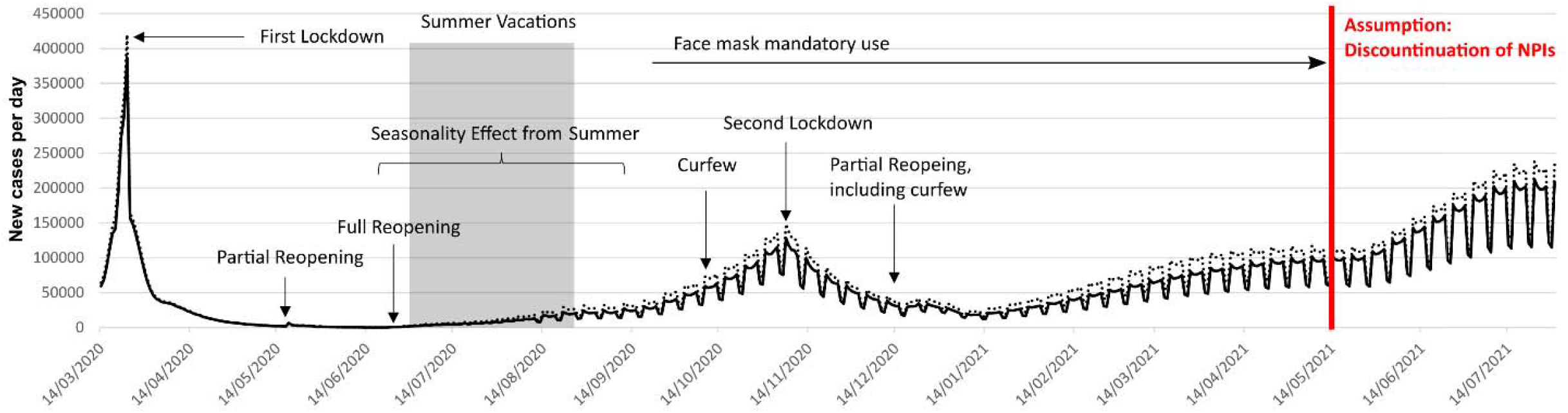
Model-predicted daily incidence in France in the scenario considering that NPIs present on December 15^th^would be maintained until April 15^th^ and then discontinued after that date. The dotted lines represent the uncertainty range (95% prediction interval) stemming from the uncertainty in the parameter values.

**eTable 1.** Summary of main model parameters.

| Parameters | Value | Source |
| --- | --- | --- |
| **Individuals' characteristics** |  |  |
| Family structure (%) |  | Insee, 2020<sup>1</sup> |
| Singles | 37% |  |
| Couples with children | 27% |  |
| Couples without children | 27% |  |
| Singles with children | 9% |  |
| Age structure (categorized by 5-year age groups) |  | Insee, 2020<sup>1</sup> |
| Prevalence of conditions associated with increased risk of death from Covid-19 (i.e., obesity, diabetes, chronic cardiac diseases, and chronic respiratory diseases) | Estimates per 10-year age groups and sex | 2-7 |
| **Social contacts** |  |  |
| School class size (average) | 30 | Assumption |
| Proportion of small companies (<10 employees) | 18.40% | INSEE, 2018<sup>8</sup> |
| Number of colleagues in small companies (average) | 2 | Assumption |
| Number of colleagues in bigger companies (average) | 10 | Assumption |
| Employment rate (for people aged 20 to 65 years) | 92% | INSEE, 2020<sup>8</sup> |
| Shopping density (per 100,000 inhabitants) | 29.3 | APUR, 2018<sup>9</sup> |
| Number of shopping trips (average per week) | 1.2 | Assumption |
| Number of people met per shopping trip (average) | 5 | Assumption |
| Social network distance | 22 | 10 |
| Frequency of meeting friends (average per week) | 1 | Assumption |
| Event participations, i.e., museum, cinema, music and sport events (average per year) | 5.4 | French Ministry of Culture, 2015<sup>11</sup> |
| Close encounters per event participation (average) | 5 | Assumption |
| Round trips with public transport (average per week) for workers | 5 | Assumption |
| Round trips with public transport (average per week) for non-workers | 1.7 | Assumption |
| Close encounters in public transport | 3 to 5 | Assumption, with work-related trips assumed to happen at peak times with more encounters |
| International contamination (average, per week) | 1.8 | Based on imported cases observed in France initially (Santé Publique France)<sup>7</sup> |
| **SARS-CoV-2 infection characteristics** |  |  |
| Contamination risk (per min/m<sup>2</sup>) | 0.025 | Estimated through model calibration, and based on |
| Proportion of the population infected with SARS-COV-2 in May 2020 | 4.5% | seroprevalence in May 2020 in France <sup>12</sup> |
| Proportion of asymptomatic people | 25% | London Imperial College, 2020 <sup>13</sup> |
| Hospitalization rates | [0.1%-31.4%] | Institut Pasteur <sup>14</sup> |
| ICU rates (if hospitalized) | [3.4%-36.4%] | Institut Pasteur <sup>14</sup> |
| Mortality rates (if hospitalized) | [0.0%-42.0%] | Institut Pasteur <sup>14</sup> |
| Impact of comorbidities on outcomes (Hospitalization, ICU, Mortality) | Hazard ratio estimates adjusted for sex and age and other comorbidities | <sup>15</sup> |
| % of Reduction of contamination risk (per min/m<sup>2</sup>) during the summer (seasonality assumption) | 23% | Kissler, 2020 <sup>16</sup> |
| **Delays (days)** |  |  |
| Incubation time (average, standard deviation) | 6.4 (2.3) | <sup>17</sup> |
| Infection onset to diagnosis (average, standard deviation) | 2.1 (2.6) | <sup>18</sup> |
| Infection onset to hospital admission (average, standard deviation) | 5.8 (4.2) | Institut Pasteur <sup>14</sup> |
| Hospital admission to recovery or transfer to rehabilitation care (average, standard deviation) (no oxygen or low-flow oxygen) | 8.0 (1.0) | DRESS <sup>19</sup> |
| Hospital admission to death (average, standard deviation) (no ICU admissions) | 4.0 (1.0) | DRESS <sup>19</sup> |
| Rehabilitation care to Recovery (average, standard deviation) (no oxygen or low-flow oxygen) | 33.0 (3.3) | DRESS <sup>19</sup> |
| Hospital admission to ICU (average, standard deviation) (initially low-flow oxygen) | 4.0 (1.0) | DRESS <sup>19</sup> |
| ICU admission to ICU discharge or death (average, standard deviation) (short stay) | 16.0 (1.6) | DRESS <sup>19</sup> |
| ICU admission to ICU discharge or death (average, standard deviation) (long stay) | 35.0 (3.5) | DRESS <sup>19</sup> |
| ICU discharge to recovery or rehabilitation care (average, standard deviation) | 4.0 (1.0) | DRESS <sup>19</sup> |
| Infection onset to recovery (average, standard deviation) (no hospitalization) | 20.5 (6.7) | London Imperial College <sup>13</sup> |
| Proportion of hospitalized patients with mild or moderate disease with low-flow oxygen | 78.2% | Assumption |
| Proportion of hospitalized patients with mild or moderate disease requiring rehabilitation care | 12.4% | DRESS <sup>19</sup> |
| Proportion of hospitalized patients with severe disease admitted directly to the ICU | 63.0% | DRESS <sup>19</sup> |
| Proportion of hospitalized patients<br>admitted to the ICU with a long stay | 15.0% | DRESS <sup>19</sup> |

